# Usual Source of Primary Care and Preventive Care Measures in a Pandemic: A Nationwide Study in Japan

**DOI:** 10.1101/2021.09.26.21263606

**Authors:** Takuya Aoki, Yasuki Fujinuma, Masato Matsushima

## Abstract

**Objectives:** In a pandemic when there are many barriers to providing preventive care by health care workers, it is unclear whether primary care contributes to the quality of preventive care and what type of preventive care delivery is a challenge for primary care providers. This study aimed to assess multiple preventive care measures and to examine their associations with having a usual source of primary care and primary care performance during the COVID-19 pandemic in Japan.

**Design:** Nationwide cross-sectional study.

**Setting:** Japanese general adult population.

**Participants:** 1,757 adult residents.

**Primary outcome measures:** Fourteen preventive care measures aggregated the overall, screening, immunization, and counseling composites.

**Results:** Depression screening, zoster vaccination, and tetanus vaccination had low implementation rates even among participants with a usual source of primary care. After adjustment for possible confounders, having a usual source of primary care was positively associated with all preventive care composites. Primary care performance assessed by the Japanese version of Primary Care Assessment Tool Short Form was also dose-dependently associated with an increase in all composites. Results of the sensitivity analyses using a different calculation of preventive care composite were similar to those of the primary analyses.

**Conclusions:** Receipt of primary care, particularly high-quality primary care, contributed to increased preventive care utilization even during the COVID-19 pandemic. However, the rate of mental health screening in primary care was at a very low level. Therefore, addressing mental health issues should be a major challenge for primary care providers during and after the pandemic.

## Introduction

Primary care has been considered to contribute to better population health, efficiency, and equity.^1,2^ This is the reason countries have raised the issue of strengthening primary care systems. For instance, a new consensus report by the National Academies of Sciences, Engineering, and Medicine emphasized that the US should prioritize the implementation of high-quality primary care by the government and private sector.^3^ In Japan, the Ministry of Health, Labour and Welfare has recommended that all individuals should have a usual source of primary care and identified the improvement of primary care performance as an important issue.^4^ In Japan, physicians trained in an internal medicine-based residency program have typically delivered primary care. In addition to them, the Japan Primary Care Association has certified family physicians since 2010^5^ and the Japanese Medical Specialty Board started a new certification program for primary care specialists in 2018.^6^

Preventive care is one of the mechanisms for the beneficial impact of primary care on population health.^1^ Several studies before the COVID-19 pandemic have examined the value of primary care in the quality of preventive care at the individual level. For example, previous studies conducted in the US reported that having a usual source of primary care is associated with better quality of preventive care.^7,8^ Other studies have demonstrated that higher levels of primary care attributes are associated with increased utilization of preventive care services.^9,10^

However, the provision of preventive care has been disrupted due to the COVID-19 pandemic. A steep decline in the utilization of preventive services, such as cancer screening and counseling, was reported in 2020. ^11,12^ During the pandemic, health care workers and facilities allocated resources to address the influx of patients with COVID-19. Government restrictions on movement and non-essential activities could be barriers to health care accessibility. Studies conducted in the US and Japan have consistently reported that the number of outpatient visits decreased while that of telemedicine visits increased during the pandemic.^13,14^ A study in the US also indicated that primary care physicians are less likely to deliver preventive care, such as blood pressure and cholesterol level assessments, during telemedicine visits compared with office-based ones.^13^

In a pandemic when there are many barriers to providing preventive care by health care workers, it becomes unclear whether primary care contributes to the quality of preventive care and what type of preventive care delivery is a challenge for primary care providers. Answering these questions is fundamental to rethinking the role of primary care during and after the COVID-19 pandemic. Therefore, in the present study, we aimed to assess multiple preventive care measures and to examine their associations with having a usual source of primary care and primary care performance during the pandemic. We used a representative sample of the Japanese general adult population.

## Methods

### Design, setting, and participants

The data for this study was sourced from the National Usual source of Care Survey (NUCS), which was conducted in May 2021. The NUCS was a nationwide mail survey that collected data on the usual source of primary care, health care utilization, health conditions, health-related quality of life, and sociodemographic characteristics of a representative sample of the Japanese adult population. In the NUCS, a nationally representative panel in Japan, which was administered by the Nippon Research Center, was used to select potential participants. This panel comprised approximately 70,000 residents who were selected using a multistage sampling method and participated in a previous survey of the Nippon Research Center.^15^ From the panel, 2,000 potential participants aged 20–75 years were selected using stratified sampling by age, sex, and residential area. The survey participants received JPY500 gift certificates.

The institutional review board of the Jikei University School of Medicine provided ethical approval for this study (approval no. 32-416(10505)).

## Measures

### Usual source of primary care

To identify an individual’s usual source of primary care, the following item was used in the Primary Care Assessment Tool (PCAT)^16^ and nationwide surveys, such as the Medical Expenditure Panel Survey (MEPS)^17^: “Is there a doctor that you usually go to if you are sick or need advice about your health?”. A participant was considered to have a usual source of primary care if he or she was able to identify a physician who practices outside of university hospitals.

For participants who have a usual source of primary care, we conducted a primary care performance assessment based on patient experience using the Japanese version of Primary Care Assessment Tool Short Form (JPCAT-SF).^18^ The JPCAT-SF is based on the PCAT^16^, which was developed by the Johns Hopkins Primary Care Policy Center. This tool is a Japanese version of the PCAT and not a simple Japanese translation of the PCAT. It consists of fewer items than the original version for better usability. A previous study showed that the JPCAT-SF has good reliability and validity.^18^ This 13-item tool comprises six multi-item subscales addressing the following primary care attributes: first contact, longitudinality, coordination, comprehensiveness (services available), comprehensiveness (services provided), and community orientation. The JPCAT-SF’s scoring system is structured as follows: each response on a 5-point Likert scale (1 = strongly disagree, 2 = somewhat disagree, 3 = not sure, 4 = somewhat agree, and 5 = strongly agree) is converted into an item score between 0 and 4. The calculated means of item scores in the same subscale are multiplied by 25 to yield subscale scores ranging from 0 to 100 points. The JPCAT-SF score is the mean of the six subscale scores and reflects an overall measure of primary care performance, with higher scores indicating better performance.

### Preventive care measures

The outcome measures in this study were defined as selected multiple preventive care measures according to the recommendations of the U.S. Preventive Services Task Force^19^ (Grade A and B) and Centers for Disease Control and Prevention.^20^ From the recommendations, we excluded measures of preventive therapies, those that had not been formally approved in Japan, and those that could not be accurately assessed using a self-administered questionnaire (e.g., measures that require an assessment of additional risk factors). After applying the exclusion criteria, we included 14 preventive care measures: colorectal cancer screening, breast cancer screening, cervical cancer screening, hypertension screening, abnormal blood glucose screening, osteoporosis screening, depression screening, influenza vaccination, pneumococcal vaccination, zoster vaccination, tetanus vaccination, smoking cessation counseling, alcohol use counseling, and weight loss counseling (Table 1). We constructed an overall preventive care composite and three clinically meaningful composites (screening, immunization, and counseling composites). The primary outcome measure in this study was the overall preventive care composite, and the secondary outcome measures were screening, immunization, and counseling composites.

**Table 1.**
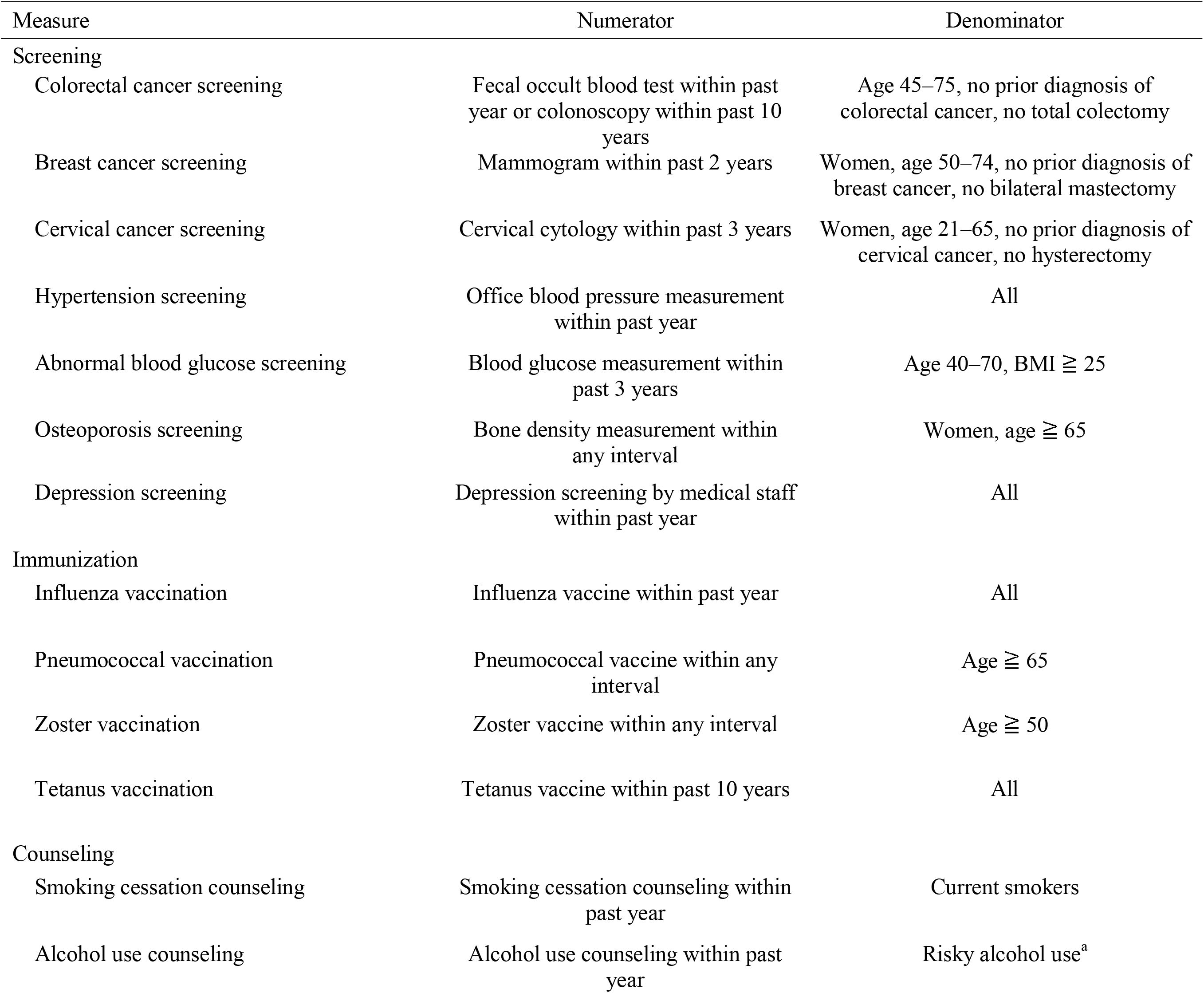
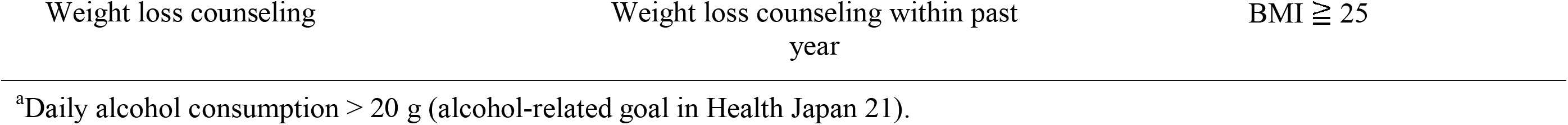
Definition of Preventive Care Measures.

To calculate each outcome measure, we first identified eligible participants for the measure and then determined whether or not they received particular preventive care. To calculate composites, we divided all instances in which the recommended service was delivered by the number of times participants were eligible for the service in the category, as others have done.^8,21^ Composites could range from 0 to 100%.

### Statistical analysis

We computed the descriptive statistics for individual preventive care measures and composites with or without a usual source of primary care. To examine whether having a usual source of primary care was associated with preventive care composites, we used multivariable linear regression analyses. The following potential confounders were included in the analyses: age, sex, marital status, years of education, employment status, annual household income, smoking status, body mass index, health literacy assessed by the Communicative and Critical Health Literacy (CCHL)^22^, number of chronic conditions, and health-related quality of life assessed by the five-level version of the EuroQol five-dimensional questionnaire (EQ-5D-5L).^23^ We used a validated list of 20 chronic conditions that were created based on previous multimorbidity literature and their relevance to the primary care population^24^: hypertension, depression/anxiety, chronic musculoskeletal conditions causing pain or limitation, arthritis/rheumatoid arthritis, osteoporosis, chronic respiratory disease (asthma, chronic obstructive pulmonary disease, or chronic bronchitis), cardiovascular disease, heart failure, stroke/transient ischemic attack, stomach problem, colon problem, chronic hepatitis, diabetes, thyroid disorder, any cancer in the previous five years, kidney disease/failure, chronic urinary problem, dementia/Alzheimer’s disease, hyperlipidemia, and obesity. All confounders were evaluated using a self-administered questionnaire. In addition, to examine the dose-response association between primary care performance and preventive care composites, we performed analyses of the outcomes in relation to the JPCAT-SF score quartile, adjusting for the same potential confounders.

We also conducted sensitivity analyses using a different calculation of the overall preventive care composite. In the sensitivity analyses, we included only measures with an interval of one year or less: colorectal cancer screening, hypertension screening, depression screening, influenza vaccination, smoking cessation counseling, alcohol use counseling, and weight loss counseling. Because participants may have received services before the pandemic for the preventive care measures with longer recommended intervals.

For each analysis, we used a two-sided significance level of *P* = 0.05, without adjustment for multiple comparisons.^25^ For missing independent variables in the regression model, we performed complete case analyses. Statistical analyses were conducted using R, version 4.1.0 (R Foundation for Statistical Computing, Vienna, Austria; www.R-project.org).

### Patient and public involvement

No patient involved.

## Results

### Participants’ characteristics

Of the 2,000 adult residents, 1,757 responded to the NUCS (response rate: 87.9%). Table 2 shows the characteristics of the study population, with or without a usual source of primary care. Among the participants, 1,011 (57.5%) had a usual source of primary care. Compared with participants without a usual source of primary care, those with a usual source of primary care were older (mean age, 53.1 vs. 45.9 years), more often female (53.9% vs. 47.3%), more frequently unemployed (29.7% vs. 20.2%), and had more chronic conditions (with ≥ 2 chronic conditions, 34.5% vs. 11.9%).

**Table 2.** Participants’ Characteristics With or Without Usual Source of Primary Care.

| Characteristic | Total<br>(N = 1,757) | Has Usual Source of<br>Primary Care<br>(n = 1,011) | No Usual Source of Primary<br>Care<br>(n = 746) |
| --- | --- | --- | --- |
| Age, mean (SD), y | 50.1 (15.1) | 53.1 (15.1) | 45.9 (14.1) |
| Gender, n (%) |  |  |  |
| Female | 898 (51.1) | 545 (53.9) | 353 (47.3) |
| Marital status, n (%) |  |  |  |
| Married | 1,333 (75.9) | 784 (77.5) | 549 (73.6) |
| Widowed | 65 (3.7) | 45 (4.5) | 20 (2.7) |
| Annulled, divorced, separated | 89 (5.1) | 55 (5.4) | 34 (4.6) |
| Never married | 269 (15.3) | 127 (12.6) | 142 (19.0) |
| Data missing | 1 (0.1) | 0 (0.0) | 1 (0.1) |
| Education, n (%) |  |  |  |
| Less than high school | 57 (3.2) | 37 (3.7) | 20 (2.7) |
| High school | 584 (33.2) | 348 (34.4) | 236 (31.6) |
| Junior college | 484 (27.5) | 279 (27.6) | 205 (27.5) |
| More than or equal to college | 590 (33.6) | 323 (31.9) | 267 (35.8) |
| Data missing | 42 (2.4) | 24 (2.4) | 18 (2.4) |
| Employment status, n (%) |  |  |  |
| Full-time employee | 691 (39.3) | 347 (34.3) | 344 (46.1) |
| Part-time employee | 387 (22.0) | 231 (22.8) | 156 (20.9) |
| Self employee | 227 (12.9) | 132 (13.1) | 95 (12.7) |
| Unemployed | 451 (25.7) | 300 (29.7) | 151 (20.2) |
| Data missing | 1 (0.1) | 1 (0.1) | 0 (0.0) |
| Annual household income, n (%), million JPY |  |  |  |
| < 3.00 (≒27,000 US dollar) | 288 (16.4) | 170 (16.8) | 118 (15.8) |
| 3.00–4.99 | 532 (30.3) | 332 (32.8) | 200 (26.8) |
| 5.00–6.99 | 435 (24.8) | 256 (25.3) | 179 (24.0) |
| 7.00–9.99 | 312 (17.8) | 167 (16.5) | 145 (19.4) |
| ≥ 10.00 | 170 (9.7) | 76 (7.5) | 94 (12.6) |
| Data missing | 20 (1.1) | 10 (1.0) | 10 (1.3) |
| Currently smoke, n (%) | 265 (15.1) | 148 (14.6) | 117 (15.7) |
| Data missing | 5 (0.3) | 5 (0.5) | 0 (0.0) |
| BMI, mean (SD) | 22.9 (3.7) | 23.2 (3.6) | 22.6 (3.7) |
| Data missing, n (%) | 10 (0.6) | 8 (0.8) | 2 (0.3) |
| CCHL, mean (SD) | 3.5 (0.7) | 3.5 (0.7) | 3.5 (0.7) |
| Data missing, n (%) | 8 (0.5) | 3 (0.3) | 5 (0.7) |
| Number of chronic conditions, n (%) |  |  |  |
| 0 | 794 (45.2) | 324 (32.0) | 470 (63.0) |
| 1 | 454 (25.8) | 297 (29.4) | 157 (21.0) |
| ≥ 2 | 438 (24.9) | 349 (34.5) | 89 (11.9) |
| Data missing | 71 (4.0) | 41 (4.1) | 30 (4.0) |
| EQ-5D-5L, mean (SD) | 0.89 (0.08) | 0.88 (0.09) | 0.90 (0.07) |
| Data missing, n (%) | 7 (0.4) | 2 (0.2) | 5 (0.7) |

Abbreviations: BMI, body mass index, CCHL, Communicative and Critical Health Literacy; EQ-5D-5L, five-level version of the EuroQol five-dimensional questionnaire.

### Preventive care measures with or without a usual source of primary care

Table 3 shows the preventive care measures in the two groups, namely, with or without a usual source of primary care. In both groups, the highest implementation rate was observed in hypertension screening (87.8% for with and 70.6% for without), and the lowest implementation rate was observed in zoster vaccination (1.9% for with and 2.5% for without). Tetanus vaccination and depression screening also had low implementation rates for both groups (tetanus vaccination, 5.2% for with and 3.6% for without; depression screening, 11.2% for with and 7.8%, for without). Having a usual source of primary care was positively associated with increased receipt of each preventive care measure, except for zoster vaccination. Table 3 also shows the adjusted associations between having a usual source of care and preventive care composites. Participants with a usual source of primary care had a higher overall composite compared with those without (mean, 43.9% vs. 33.9%; adjusted mean difference, 7.2% [95% CI, 5.2% to 9.1%]). Having a usual source of primary care was significantly associated with an increase in all composites. The largest difference was observed in counseling composite (adjusted mean difference, 8.0% [95% CI, 1.6% to 14.3%]).

**Table 3.** Preventive Care Measures With or Without Usual Source of Primary Care.

| Measure | Has Usual Source of Primary Care<br>(n = 1,011) |  | No Usual Source of Primary Care<br>(n = 746) |  | Adjusted Mean<br>Difference<br>(95%CI)<sup>a</sup> | P value |
| --- | --- | --- | --- | --- | --- | --- |
|  | n | Mean, % (95%CI) | n | Mean, % (95%CI) |  |  |
| Overall composite | 1,011 | 43.9 (42.7 to 45.1) | 746 | 33.9 (32.4 to 35.3) | 7.2 (5.2 to 9.1) | <0.001 |
| Screening composite | 1,011 | 56.3 (54.8 to 57.8) | 746 | 45.0 (43.0 to 47.0) | 7.0 (4.4 to 9.6) | <0.001 |
| Colorectal cancer screening | 657 | 67.1 (63.5 to 70.7) | 375 | 51.5 (46.4 to 56.5) |  |  |
| Breast cancer screening | 296 | 51.0 (45.3 to 56.7) | 130 | 50.8 (42.1 to 59.5) |  |  |
| Cervical cancer screening | 357 | 62.5 (57.4 to 67.5) | 298 | 58.1 (52.4 to 63.7) |  |  |
| Hypertension screening | 1,011 | 87.8 (85.8 to 89.9) | 746 | 70.6 (67.4 to 73.9) |  |  |
| Abnormal blood glucose screening | 190 | 84.2 (79.0 to 89.4) | 99 | 69.7 (60.5 to 78.9) |  |  |
| Osteoporosis screening | 165 | 77.6 (71.1 to 84.0) | 44 | 63.6 (48.8 to 78.4) |  |  |
| Depression screening | 1,011 | 11.2 (9.2 to 13.1) | 746 | 7.8 (5.8 to 9.7) |  |  |
| Immunization composite | 1,011 | 28.6 (27.1 to 30.1) | 746 | 20.8 (19.1 to 22.6) | 7.9 (5.4 to 10.3) | <0.001 |
| Influenza vaccination | 1,011 | 59.8 (56.8 to 62.9) | 746 | 41.6 (38.0 to 45.1) |  |  |
| Pneumococcal vaccination | 305 | 55.4 (49.8 to 61.0) | 93 | 44.1 (33.8 to 54.4) |  |  |
| Zoster vaccination | 616 | 1.9 (0.9 to 3.0) | 282 | 2.5 (0.7 to 4.3) |  |  |
| Tetanus vaccination | 1,011 | 5.2 (3.9 to 6.6) | 746 | 3.6 (2.3 to 5.0) |  |  |
| Counseling composite | 494 | 44.5 (40.4 to 48.6) | 338 | 26.9 (22.4 to 31.5) | 8.0 (1.6 to 14.3) | 0.014 |
| Smoking cessation counseling | 148 | 46.6 (38.5 to 54.8) | 117 | 25.6 (17.6 to 33.7) |  |  |
| Alcohol use counseling | 226 | 24.8 (19.1 to 30.5) | 176 | 19.9 (13.9 to 25.8) |  |  |
| Weight loss counseling | 276 | 62.0 (56.2 to 67.7) | 154 | 38.3 (30.5 to 46.1) |  |  |

<sup>a</sup>Adjusted for age, sex, marital status, years of education, employment status, annual household income, smoking status, BMI, health literacy, number of chronic conditions, and EQ-5D-5L; positive difference, participants with primary care received more preventive service.

### Primary care performance and preventive care measures

Table 4 shows the associations between primary care performance, assessed by the JPCAT-SF, and preventive care composites. Primary care performance was dose-dependently associated with an increase in all composites, including the overall composite (adjusted mean difference, 9.9% [95% CI, 7.0% to 12.9%] for the JPCAT-SF highest quartile, compared with no usual source of primary care). The largest association was observed in the counseling composite (adjusted mean difference, 17.0% [95% CI, 7.8% to 26.2%] for the JPCAT-SF highest quartile, compared with no usual source of primary care).

**Table 4.**
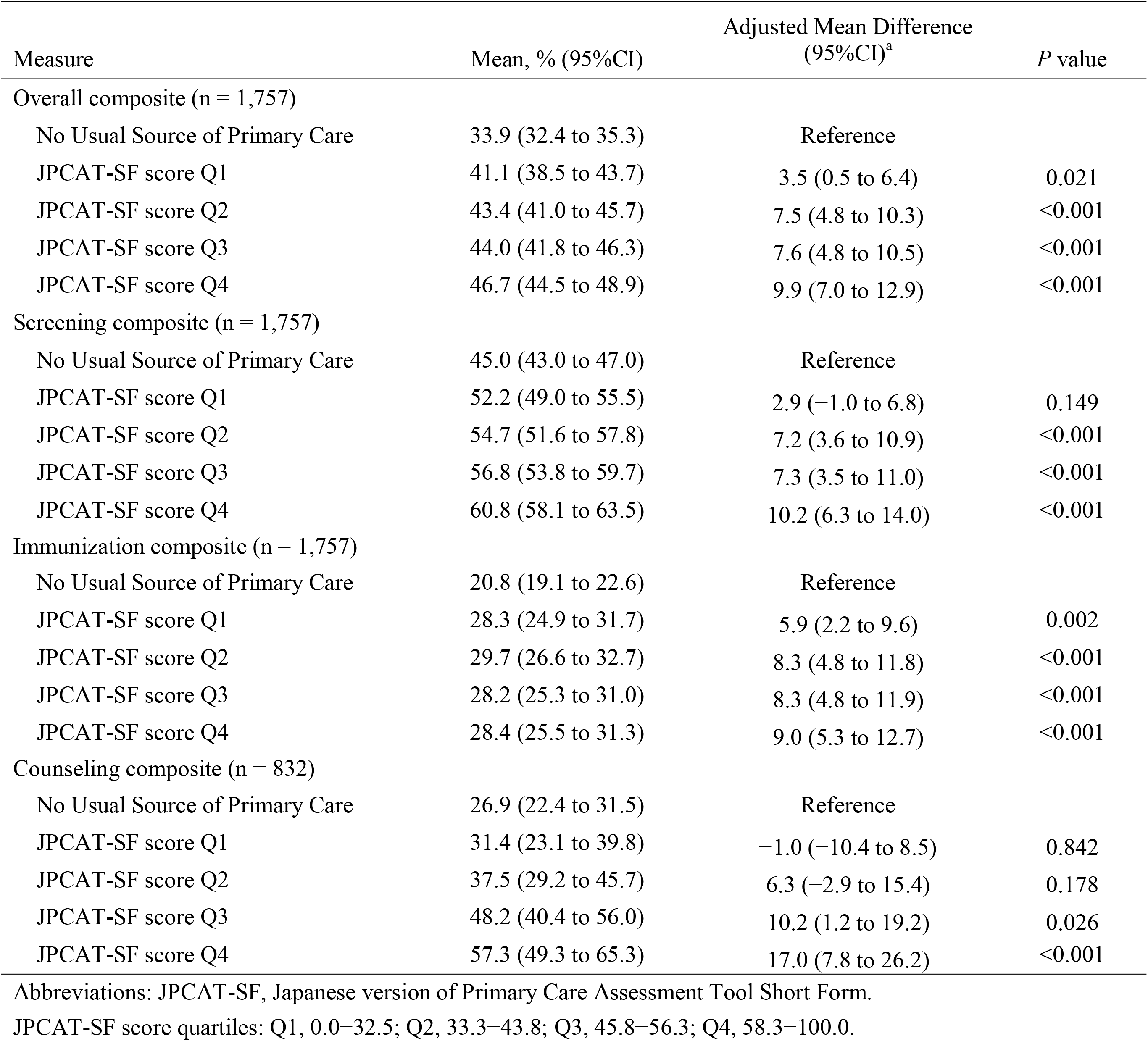
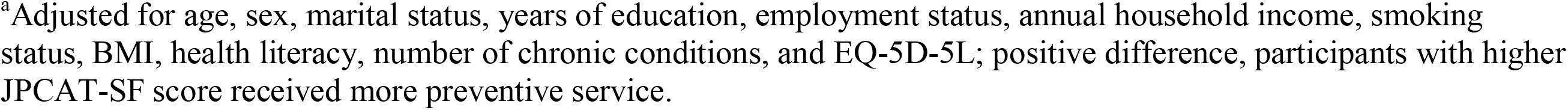
Associations Between Primary Care Performance and Preventive Care Measures.

Table 5 shows the results of the sensitivity analyses using different calculations of the overall preventive care composite (including only measures with an interval of one year or less). The findings are similar to those in the primary analyses, indicating that having a usual source of primary care and primary care performance being positively associated with the overall composite.

**Table 5.** Sensitivity Analyses for Overall Preventive Care Composite (Including Only Measures With Interval of One Year or Less)^a^.

|  | Overall composite<br>Mean, % (95% CI) | Adjusted Mean Difference<br>(95% CI)<sup>b</sup> | *P* value |
| --- | --- | --- | --- |
| No Usual Source of Primary Care | 39.4 (37.6 to 41.3) | Reference |  |
| Has Usual Source of Primary Care | 53.7 (52.3 to 55.2) | 10.2 (7.7 to 12.6) | <0.001 |
| No Usual Source of Primary Care | 39.4 (37.6 to 41.3) | Reference |  |
| JPCAT-SF score Q1 | 49.9 (46.6 to 53.1) | 5.6 (1.9 to 9.3) | 0.003 |
| JPCAT-SF score Q2 | 51.6 (48.7 to 54.5) | 9.9 (6.5 to 13.4) | <0.001 |
| JPCAT-SF score Q3 | 54.2 (51.4 to 57.0) | 10.8 (7.2 to 14.3) | <0.001 |
| JPCAT-SF score Q4 | 58.7 (55.9 to 61.6) | 14.2 (10.5 to 17.8) | <0.001 |

Abbreviations: JPCAT-SF, Japanese version of Primary Care Assessment Tool Short Form.
JPCAT-SF score quartiles: Q1, 0.0–32.5; Q2, 33.3–43.8; Q3, 45.8–56.3; Q4, 58.3–100.0.
<sup>a</sup>Included colorectal cancer screening, hypertension screening, depression screening, influenza vaccination, smoking cessation counseling, alcohol use counseling, and weight loss counseling.
<sup>b</sup>Adjusted for age, sex, marital status, years of education, employment status, annual household income, smoking status, BMI, health literacy, number of chronic conditions, and EQ-5D-5L; positive difference, participants with higher JPCAT-SF score received more preventive service.

## Discussion

Our nationwide study of the Japanese adult population revealed that having a usual source of care was positively associated with multiple preventive care measures, including screening, immunization, and counseling during the COVID-19 pandemic. Our study also found that primary care performance was dose-dependently associated with increased receipt of preventive care. These findings indicate that receipt of primary care, particularly high-quality primary care, contributes to increased preventive care even during a pandemic when there are many barriers to providing preventive care by health care workers.

To the best of our knowledge, this is the first study to report the contribution of primary care to multiple preventive care measures during a pandemic. Our findings are consistent with prior studies before the pandemic, showing that having a usual source of primary care and primary care performance is positively associated with receipt of preventive care.^7-10,26,27^ This association has been unknown since the pandemic; therefore, this study expanded the evidence of the value of primary care in preventive care during a pandemic or health care crisis. Primary care attributes, such as first contact, longitudinality, coordination, and comprehensiveness, which are essential to high-performance primary care, may be effective in improving population health through better quality of preventive care, even during and after the pandemic. Policymakers and health care system leaders in Japan should consider implementing a patient registration system to ensure that all residents have a usual source of primary care and strongly promote the training of certified primary care specialists for high-quality primary care.

However, we found the implementation rates of depression screening and some immunizations that are not related to respiratory infections to be at very low levels, even among participants with a usual source of primary care. Especially, depression screening is a crucial preventive care measure because the number of residents suffering from mental health problems has increased due to the pandemic.^28^ The low rate of depression screening in primary care settings has been a problem before the pandemic.^29^ During the pandemic, a psychological assessment in primary care should be promoted and include queries about pandemic-related stressors, secondary adversities (e.g., economic loss), and psychosocial effects (e.g., substance use and domestic violence).^30^ Addressing mental health issues should be a major challenge for primary care providers during and after the pandemic.

A key strength of our study is the use of data from a nationwide study, with a sample representative of the Japanese adult population, which allows for generalization of its results to the wider population. Another strength is the high study response rate compared with other national surveys, such as the MEPS, which has often been used to investigate the association between receipt of primary care and the quality of care. The PCAT is a validated and internationally established tool for evaluating the performance of primary care attributes. In multivariable analyses, we adjusted for important potential confounders, including health literacy, chronic conditions, and health-related quality of life.

However, the present study has several limitations. First, our outcome measures did not address all preventive care qualities. For example, we excluded measures of preventive therapies and those that could not be assessed accurately using a questionnaire. Second, although a self-reported survey is a useful method for evaluating preventive care measures in a national study, social desirability and recall biases could have affected our results by overestimating the measures and the associations of interest. Third, for preventive care measures with longer recommended intervals, such as tetanus vaccination, participants’ usual source of primary care might have changed in the interval between receipt of preventive care and study participation. Fourth, given that the data were cross-sectional, a causal relationship between receipt of primary care and preventive care measures cannot be established definitively.

## Conclusion

Our nationwide study of the Japanese adult population revealed that receipt of primary care, particularly high-quality primary care, contributed to an increase in preventive care utilization even during the COVID-19 pandemic when there are many barriers to providing preventive care by health care workers. However, the rate of mental health screening in primary care was at a very low level. Therefore, addressing mental health issues should be a major challenge for primary care providers during and after the pandemic.

## Data Availability

Due to the nature of this research, participants of this study did not agree for their data to be shared publicly, so supporting data is not available.

## Acknowledgments

None.

## Contributors

All authors (TA, YF, and MM) of the paper contributed the conception or design of the work. TA performed the statistical analyses. TA, YF, and MM interpreted the analyses. TA drafted the manuscript. All authors reviewed and edited the manuscript, contributed to the discussion of the data and performed critical review of the manuscript. All authors gave the final approval of the manuscript before submission.

